# Will an imperfect vaccine curtail the COVID-19 pandemic in the U.S.?

**DOI:** 10.1101/2020.05.10.20097428

**Authors:** Enahoro Iboi, Calistus N. Ngonghala, Abba B. Gumel

## Abstract

The novel coronavirus (COVID-19) that emerged from Wuhan city of China in late December 2019 continue to pose devastating public health and economic challenges across the world. Although the community-wide implementation of basic non-pharmaceutical intervention measures, such as social-distancing, quarantine of suspected COVID-19 cases, isolation of confirmed cases, use of face masks in public, and contact-tracing, have been quite effective in curtailing and mitigating the burden of the pandemic, it is universally believed that the use of an anti-COVID-19 vaccine is necessary to build the community herd immunity needed to effectively control and eliminate the pandemic. This study is based on the design and use of a mathematical model for assessing the population-level impact of a hypothetical imperfect anti-COVID-19 vaccine on the control of COVID-19. An analytical expression for the minimum number of unvaccinated susceptible individuals needed to be vaccinated to achieve vaccine-induced community herd immunity is derived. The epidemiological consequence of the herd immunity threshold is that the disease can be effectively controlled or eliminated if the minimum herd immunity threshold is achieved in the community. Simulations of the model, using baseline parameter values obtained from fitting the model with mortality data relevant to COVID-19 dynamics in the US states of New York and Florida, as well as for the entire US, show that, for an anti-COVID-19 vaccine with an assumed protective efficacy of 80%, the minimum herd immunity threshold for the entire US, state of New York and state of Florida are, respectively, 90%, 84% and 85%. Furthermore, it was shown that, while a significantly large increase in vaccination rate (from baseline) is necessarily needed to eliminate COVID-19 from the entire US, the pandemic can be eliminated from the states of New York and Florida if the vaccination rate is marginally increased (by as low as 10%) from its baseline value. The prospect of COVID-19 elimination in the US or in the two states of New York and Florida is greatly enhanced if the vaccination program is combined with a public mask use program or an effective social-distancing measure. Such combination of strategies significantly reduces the vaccine-induced herd immunity threshold. Finally, it is shown that the vaccination program is more likely to lead to COVID-19 elimination in the state of Florida, followed by the state of New York and then the entire US.

## 1 Introduction

In December 2019, the Wuhan Municipal Health Commission, China, reported a cluster of a pneumonia-like illness, caused by a coronavirus (SARS-CoV) in Wuhan, Hubei Province of China [1–3]. The disease rapidly spread to many countries, resulting in the World Health Organization declaring the novel coronavirus (COVID-19) as a pandemic on March 11, 2020 [1, 2, 4]. As of May 8, 2020, COVID-19 has caused over 4 million confirmed cases and 275,000 fatalities globally. The United States, which is now the epicenter of COVID-19, recorded over 1.3 million confirmed cases and 78,500 deaths [5], with the US states of New York and Florida each recording over 340,000 and 39,000 confirmed cases and 26,500 and 1,660 deaths, respectively. While some COVID-19-infected people experience mild symptoms (such as fever, coughing and shortness of breath), others develop severe disease and complications (causing acute respiratory failure, which damages the lungs and often cause death) [4, 6, 7]. In addition to the devastating public health burden it induced in the US, COVID-19 has also inflicted severe economic hardship, causing a decrease in both human and economic productivity (translating to about 30% unemployment rate, and a projected 50% decline in GDP by the summer of 2020 [8, 9]).

There are currently no approved vaccines and effective therapies for use against COVID-19. Hence, control efforts against COVID-19 are focused on using basic non-pharmaceutical interventions (NPIs), such as social-distancing, quarantine of people suspected of being exposed to COVID-19, isolation of confirmed cases, use of face masks in public and contact-tracing [4, 10, 11]. There is now a concerted global effort to develop a safe and effective anti-COVID-19 vaccine to help combat the COVID-19 pandemic. Although the development of new vaccines generally take years, a number of pharmaceutical companies, with support from numerous government agencies and regulatory authorities, are doing everything possible to fast-track the availability of a safe and effective vaccine against COVID-19. Over a dozen organizations have announced plans to start efficacy testing for their candidate vaccines [6, 12, 13]. In fact, scientists at the Jenner Institute at the Oxford University have developed a new coronavirus vaccine that is undergoing early-phase trials, and they expect the first few million doses of their vaccine to be widely available as early as September 2020 [12, 13]. A Maryland-based biotechnology company *Novavax* has begun human clinical trials for its candidate vaccine in Australia [6]. Similarly, *Mesoblast*, a company supported by the National Institutes of Health (NIH), started a 240-patient clinical trial aimed at testing whether cells derived from bone marrow could help patients who developed a deadly immune reaction to the coronavirus [6]. Thus, there is an urgent need to develop and use mathematical models to assess the population-level impact of a future hypothetical vaccine against COVID-19. This forms the objective of the current study.

Numerous mathematical models have been developed and used to study the transmission dynamics and control of COVID-19. For instance, an agent-based model was developed by Ferguson *et al*. [10] to investigate the impact of NPIs on COVID-19-induced mortality, showing an alarming worst-case projection of number of cases for the US and Great Britain (at the tune of 81%). Eikenberry *et al*. [11] developed a multi-group model for assessing the community-wide impact of mask use by the general, asymptomatic public, a portion of which may be asymptomatically-infectious. The study shows that the use of face masks by members of the general public is potentially of high value in curtailing community transmission and the burden of the pandemic, and that the community-wide benefits are likely to be greatest when face masks are used in conjunction with other non-pharmaceutical practices (such as social-distancing), and when adoption is nearly universal (nation-wide) and compliance is high. Ngonghala *et al*. [4] developed a comprehensive model for assessing the impact of the major NPIs (quarantine, isolation, social-distancing, face mask usage in public, testing and contact-tracing) on the control of COVID-19. Their study estimated that, while the early relaxation or lifting social distancing measures and mask usage is likely to lead to second wave, the extended adherence to the current social-distancing and mask usage can significantly reduce COVID-induced mortality in the US in general, and the state of New York in particular. Mizumoto and Chowell [14] modeled the potential for a COVID-19 outbreak in the Diamond Princess cruise ship. Their study showed an estimate of a higher reproduction number, which substantially decreases with increasing effectiveness of quarantine measures on the ship. A stochastic model was developed by Hellewell *et al*. [15] to assess the impact of contact-tracing and isolation on disease control. Their study suggests that (for most instances) the spread of COVID-19 can be effectively contained in 3 months if the two control measures (contact-tracing and isolation) are highly effective. Kucharski *et al*. [16] also developed a stochastic model to assess the COVID-19 trajectory in Wuhan from January to February, 2020, showing a reduction in transmission when travel restrictions were implemented. Li *et al*. developed a mathematical model to assess the effect of mass influenza vaccination on the spread of COVID-19 and other influenza-like pathogens co-circulating during an influenza season. Their study showed that increasing influenza vaccine uptake, or enhancing the public health interventions, would facilitate the management of outbreaks of respiratory pathogens circulating during the peak flu season.

The current study is based on the design of a mathematical model to assess the impact of an anti-COVID-19 vaccine on the transmission dynamics of the COVID-19 pandemic. The model will be parametrized using available COVID-19 mortality data for the US, the state of New York, and the state of Florida to estimate important parameters related to vaccination and other control intervention (notably the parameter related to the reduction in community contacts).

## 2 Materials and Methods

### 2.1 Model Formulation

We develop a basic model for assessing the population-level impact of a potential hypothetical anti-COVID-19 vaccine. The model is developed by splitting the total human population at time t, denoted by *N*(*t*), into the mutually-exclusive compartments of unvaccinated susceptible (*S_u_*(*t*)), vaccinated susceptible (*S_v_* (*t*)), exposed (*E*(*t*)), symptomatically-infectious (*I_s_*(*t*), asymptomatically-infectious (*I_a_*(*t*)), hospitalized (*I_h_*(*t*)) and recovered (*R*(*t*)) individuals. Thus,

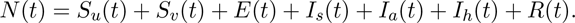

It should be emphasized that the epidemiological compartment *I_a_* also contain individuals who may show mild symptoms of the disease. Furthermore, the compartment *I_h_* for hospitalization also includes those with clinical symptoms of COVID-19 who are self-isolating at home. The model is given by the following deterministic system of nonlinear differential equations (where a dot represents differentiation with respect to time *t*):

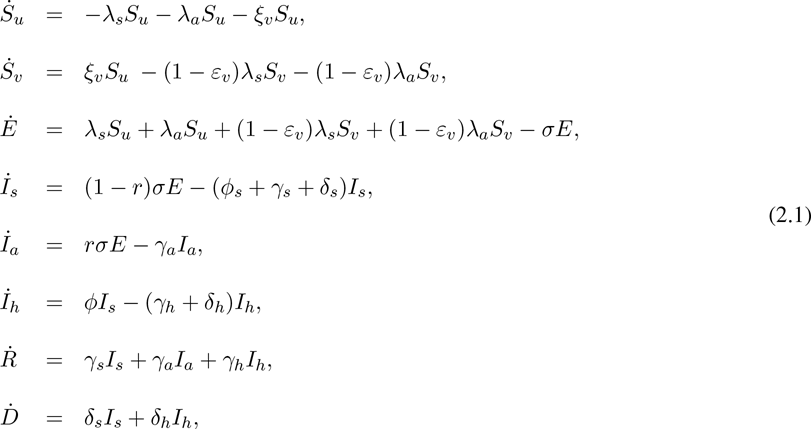

where the force of infection is given by

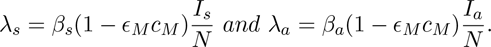

A flow diagram of the model is depicted in Figure 1. The state variables and parameters of the model are described in Tables 1 and 2, respectively. In the model (2.1), the parameter *β_s_*(*β_a_*) represents the effective contact rate (i.e., contacts capable of leading to COVID-19 transmission) for symptomatically-infectious (asymptomatically-infectious) individuals, 0 < *c_M_* ≤ 1 is the proportion of members of the public who wear face-masks (correctly and consistently) in public and 0 < *e_M_ ≤* 1 is the efficacy of the face-masks. This formulation (i.e., *β_s_* ≠ *β_a_*) allows us to account for the possible heterogeneity in the contact rates of infectious with (*I_s_*) or without (*I_a_*) clinical symptoms of COVID-19. Unvaccinated susceptible individuals are vaccinated at a *per capita* rate *ξ_v_*. It is assumed that the hypothetical anti-COVID-19 vaccine is imperfect (i.e., it allows for breakthrough infected in vaccinated susceptible individuals who became infected with COVID-19), with a protective efficacy 0 *< ε_v_ ≤* 1 against COVID-19. The parameter *σ* represents the progression rate of exposed individuals (i.e., 1/*σ* is the intrinsic incubation period of COVID-19). A proportion, 0 < *r* ≤ 1, of exposed individuals show no clinical symptoms of COVID-19 (and move to the class *I_a_*) at the end of the incubation period. The remaining proportion, 1 − *r*, show clinical symptoms and move to the *I_s_* class. The parameter *γ_s_* (*γ_a_*)(*γ_h_*) represents the recovery rate for individuals in the *I_s_* (*I_a_*)(*I_h_*) class. Similarly, *ϕ_s_* is the hospitalization (or self-isolation) rate of individuals with clinical symptoms of COVID-19. Finally, the parameter *δ_s_*(*δ_h_*) represents the COVID-induced mortality rate for individuals in the *I_s_*(*I_h_*) class. It should be mentioned that, since not much data is available on the expected properties of a future anti-COVID-19 vaccine, we do not assume that the vaccine offers any therapeutic benefits. Further, we do not assume that the vaccine wanes during the one-year time period we would choose for the numerical simulations of our model.

**Table 1:**
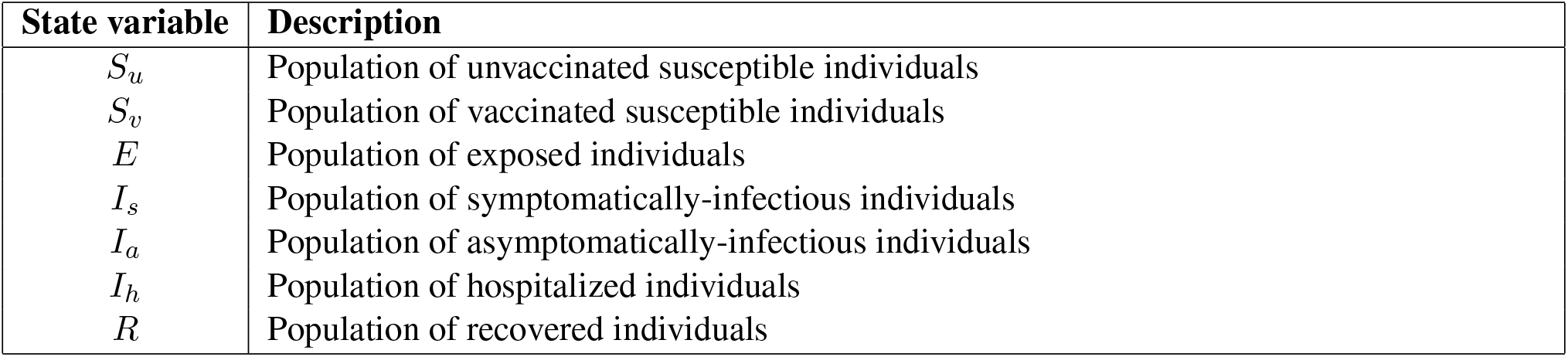
Description of state variables of the COVID-19 model.

**Table 2:**
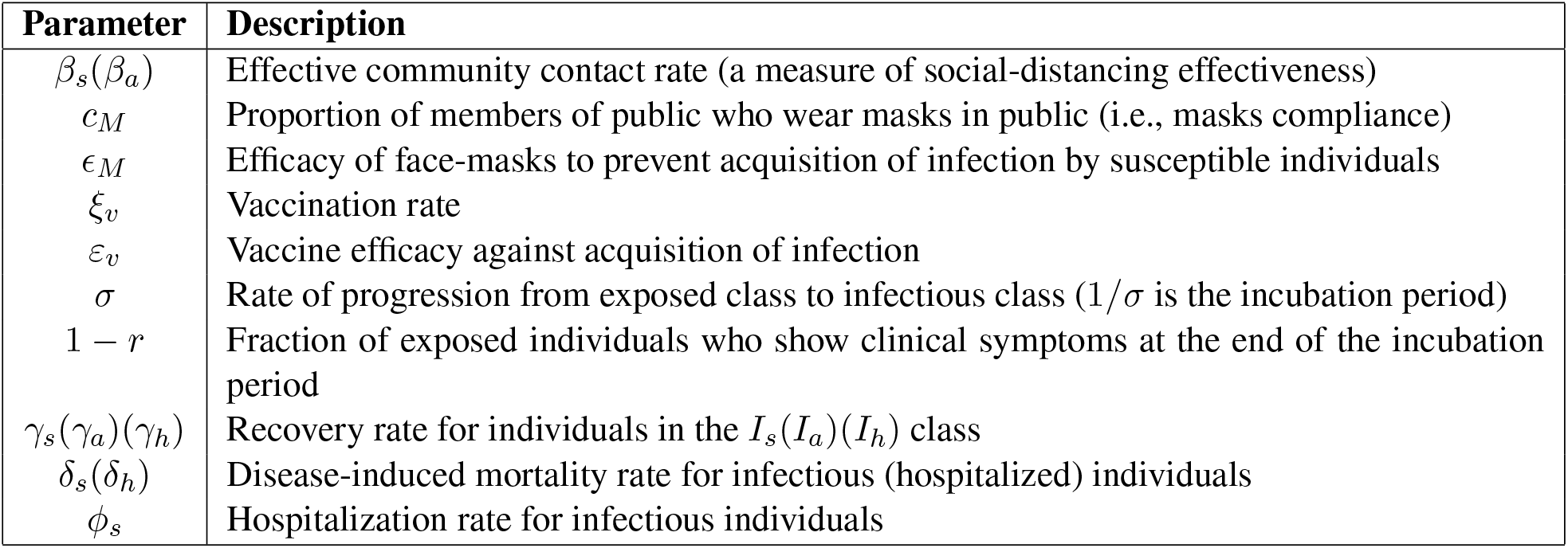
Description of the parameters of the COVID-19 model (2.1).

**Figure 1:**
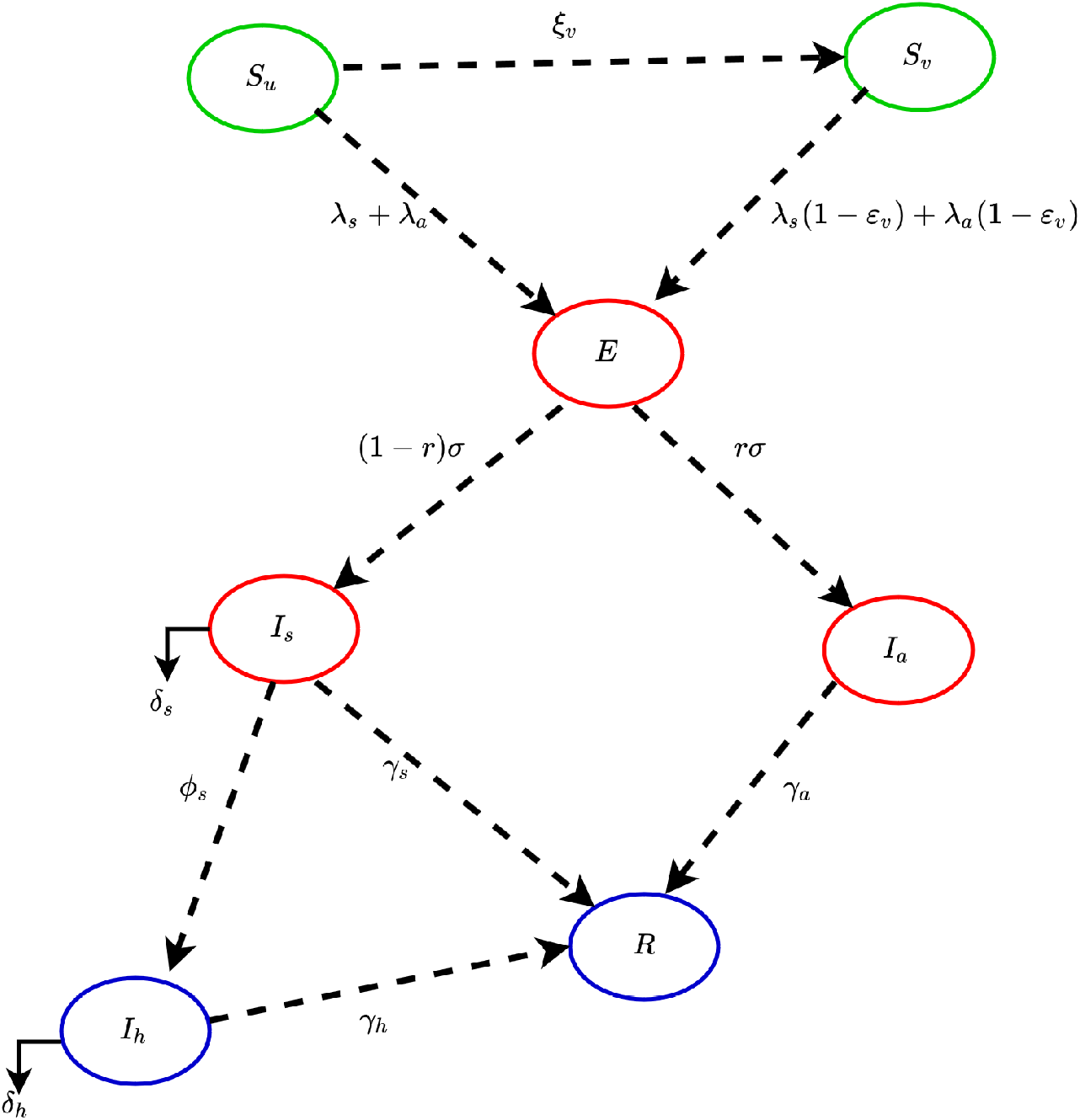
Flow diagram of the anti-COVID-19 model (2.1).

The model (2.1) is an extension of numerous deterministic models for the transmission dynamics of COVID-19 pandemic, such as those in [4, 11], by explicitly incorporating an imperfect anti-COVID-19 vaccine. To the best of our knowledge, this is the first deterministic model for COVID-19 transmission that explicitly incorporates an imperfect vaccine.

## 3 Results

### 3.1 Analytical Results: Asymptotic Stability Analysis of Continuum of Disease-free Equilibria

The model (2.1) has a continuum of disease-free equilibria (DFE), given by

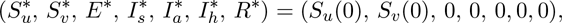

where *S_u_*(0) and *S_v_*(0) are the initial total sizes of the populations unvaccinated and vaccinated susceptible individuals, respectively (so that, *N*(0) = *S_u_*(0) + *S_v_*(0)). The next generation operator method [17, 18] can be used to analyse the asymptotic stability property of the DFE. In particular, using the notation in [17], it follows that the associated next generation matrices, *F* and *V*, for the new infection terms and the transition terms, are given, respectively, by

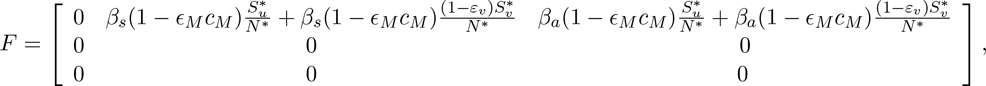

and,

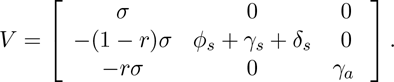

It is convenient to define the quantity 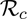 by:

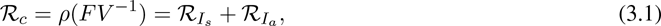

where *ρ* is the spectral radius,

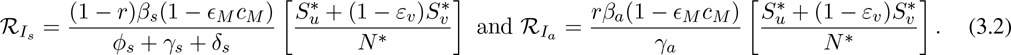

The quantity 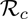 is the *control reproduction number* of the model (2.1). It measures the average number of new COVID-19 cases generated by a typical infectious individual introduced into a population where a certain fraction is protected, e.g., vaccinated (with the hypothetical vaccine) against COVID-19. It is the sum of the constituent reproduction numbers associated with the number of new cases generated by symptomatically-infectious humans (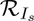) and asymptomatically-infectious (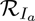) individuals. The reproduction number 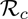 will now be interpreted epidemiologically below.

#### Interpretation of the Reproduction Number

As stated above, the reproduction number 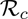 is the sum of the two constituent reproduction numbers, 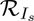 and 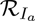. The constituent reproduction number 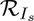 is given by the product of the infection rate of unvaccinated and vaccinated susceptible individuals by symptomatically-infectious humans, near the 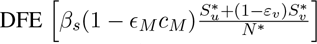, the proportion of exposed individuals that survived the incubation period and moved to the symptomatically-infectious class 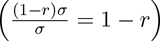 and the average duration in the asymptomatically-infectious class 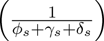. Similarly, the constituent reproduction number 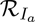 is given by the product of the infection rate of unvaccinated and vaccinated susceptible individuals, near the 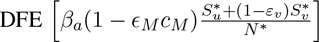, the proportion that survived the exposed class and moved to the asymptomatically-infectious class 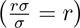 and the average duration in the *I_a_* class 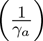. The sum of 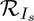 and 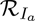 gives 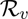.

The result below follows from Theorem 2 of [17].

##### Theorem 3.1.

*The disease-free equilibrium* (*DFE*) *of the model* (2.1) *is locally-asymptotically stable if* 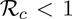*, and unstable if* 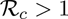.

The epidemiological implication of Theorem 3.1 is that a small influx of COVID-19 cases will not generate a COVID-19 outbreak if the control reproduction number (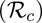) is less than unity.

The *basic reproduction number* of the model (2.1) (denoted by 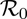), obtained by setting the control-related parameters and state variables (i.e., *c_m_*, *ε_M_*, *ε_v_* and 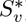) to zero in (3.1), is given by

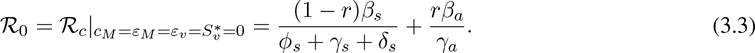

The quantity 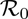 represents the average number of secondary infections generated by a typical infected individual introduced into a completely susceptible population during the duration of the infectiousness of the individual.

### 3.2 Model fitting and parameter estimation

Some of the parameters for Model (2.1) were extracted (or estimated based on COVID-19 information) from the literature (Table 3(a)), while the rest (see Table 3(b)) were estimated using mortality data for the entire US, the state of New York, and Florida [19–22]. A nonlinear least squares method was used to determine the set of parameters that minimizes the sum of the squares of the differences between the cumulative death predictions of the model (2.1) and the observed cumulative COVID-19 mortality for the entire US (Figure 2 (d)), the state of New York (Figures 2 (e)) and the state of Florida (Figures 2 (f)). Furthermore, the observed daily deaths are plotted alongside the predicted daily deaths generated using the model (2.1) with the parameter values obtained from the fitting of the cumulative cases. In particular, the plots for the observed and predicted daily deaths for the entire US, the state of New York and the state of Florida are depicted in Figures 2 (a), (b) and (c), respectively.

**Table 3:** Parameters of the model (2.1).

| (a) Parameter values from the literature |  |  | (b) Fitted parameters. |  |  |  |
| --- | --- | --- | --- | --- | --- | --- |
| Parameter | Default values | Reference | Parameter | US | New York | Florida |
| $\varepsilon_M$ | 0.5 | Estimated from [23] | $\beta_a$ | 1.2000 | 0.8069 | 1.0964 |
| $c_M$ | 0.1 | Estimated from [4] | $\beta_s$ | 0.5530 | 0.7102 | 0.6601 |
| $\varepsilon_v$ | 0.8 | Estimated | $\xi_v$ | 0.0010 | 0.0121 | 0.0144 |
| $\sigma$ | $1/5.1 \text{ day}^{-1}$ | [24, 25] | $\phi_s$ | 0.1442 | 0.0756 | 0.3716 |
| $r$ | 0.5 | [10, 25–27] | | | | |
| $\phi_s$ | $0.025 \text{ day}^{-1}$ | [10, 28] | | | | |
| $\gamma_a$ | $1/7 \text{ day}^{-1}$ | [28, 29] | | | | |
| $\gamma_s$ | $1/7 \text{ day}^{-1}$ | [28, 29] | | | | |
| $\gamma_h$ | $1/14 \text{ day}^{-1}$ | [28, 29] | | | | |
| $\delta_s$ | $0.015 \text{ day}^{-1}$ | [10] | | | | |
| $\delta_h$ | $0.0225 \text{ day}^{-1}$ | [10] | | | | |

**Figure 2:**
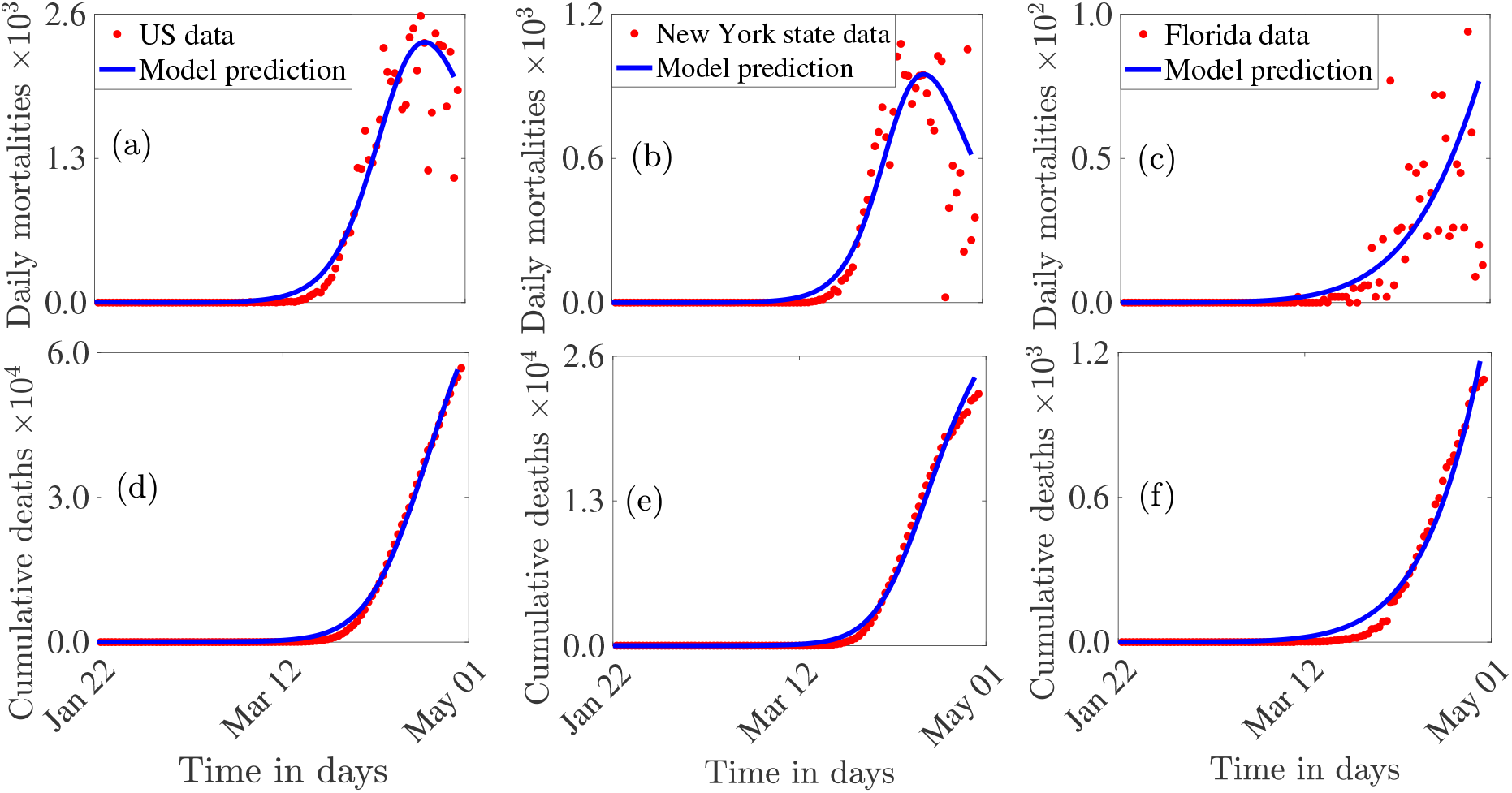
Data fitting of the model (2.1) using COVID-19 mortality data for the entire US (a) and (d), the state of New York (b) and (e), and the state of Florida (c) and (f) [19–22].

### 3.3 Numerical Simulations: Effect of Social-distancing in the Absence of Vaccination and Mask Usage

The model (2.1) is simulated, first of all, for the special case where the vaccination and public mask usage interventions are not implemented in the community. This allows for the assessment of the impact of social-distancing measures (aimed at reducing community transmission) as the sole anti-COVID-19 intervention strategy in the community. In the context of the model (2.1), social-distancing is measured in terms of the reduction in the numerical values of the associated community contact rate parameters (*β_a_* and *β_s_*) from their baseline values. The simulation results obtained, depicted in Figure 3, shows a marked decrease in disease burden (measured in terms of daily and cumulative COVID-induced mortality) with increasing effectiveness of social-distancing, as expected. For instance, while Figure 3 shows that up to 120,000 cumulative mortality can be recorded in the US if social-distancing measures were not implemented, the cumulative mortality dramatically decreases to about 64,000 (representing about 47% reduction in cumulative mortality) if strict social-distancing measures were implemented (Figure 3 (b)). Similarly, 80% and 70% reductions in cumulative mortality are, respectively, obtained for the states of New York (Figures 3 (c) and (d)) and Florida (Figures 3 (e) and (f)). These results are consistent with those reported by Ngonghala *et al*. [4], which was based on using a novel mathematical model that incorporates the main nonpharmaceutical interventions against COVID-19 (i.e., the model in [4] does not incorporate an anti-COVID-19 vaccine).

**Figure 3:**
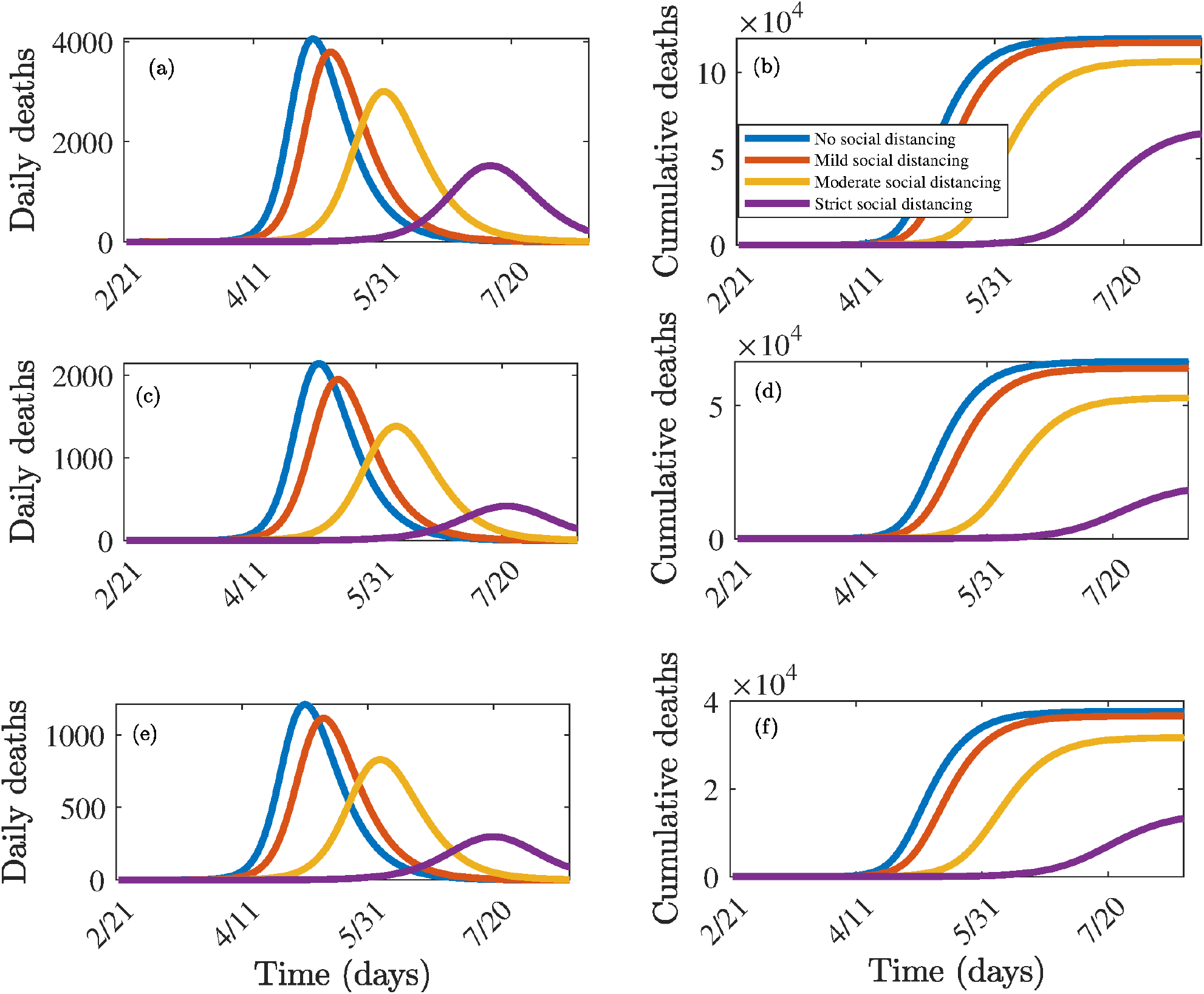
Simulations of the model (2.1) for various effectiveness of social-distancing control measures in the absence of vaccination. Mild social-distancing corresponds to 10% decrease in the values of *β_a_* and *β_s_* from their baseline values. Moderate social-distancing corresponds to 30% decrease in the values of *β_a_* and *β_s_* from their baseline values. Strict (high) social-distancing corresponds to 50% decrease in the values of *β_a_* and *β_s_* from their baseline values.(a) Daily cases for US, (b) cumulative deaths for US, (c) Daily cases for New York state, (d) cumulative deaths for New York state (e) Daily cases for Florida state, (f) cumulative deaths for Florida state. Parameter values used are as given in Table 3 (a), with various values of *β_a_* and *β_s_*.

### 3.4 Vaccine-induced Herd Immunity Threshold

For vaccine-preventable diseases, not all susceptible individuals can be immunized for various reasons, such as they are too young to be vaccinated (vaccinating infants or young children can, sometimes, harm the infants/children), they have co-morbidities and weakened immune system (and vaccinating may make their prognosis worse), they are of advanced age or they opt out for cultural, traditional or religious reasons. The question then is what is the minimum proportion of those we can vaccinate that we need to vaccinate so that those that we cannot vaccinate can be protected from developing severe disease or dying of the disease. The notion of *herd immunity* in the transmission dynamics is associated with the indirect protection, against acquisition of a communicable disease, members of the community receive when a large percentage of the population has become immune to the infectious disease due to natural recovery from prior infection or vaccination [30, 31]. The consequence of herd immunity is that individuals who are not immune (e.g., those who cannot be vaccinated or those who have not been infected yet) receive some protection against the acquisition of the disease. The safest and fastest way to achieve herd immunity is obviously vaccination. It should, however, be mentioned that Sweden employed the other mechanism for achieving herd immunity in the context of COVID-19 dynamics in Sweden [32]. In other words, the Swedish public health agencies aimed to build herd immunity by not implementing the basic community transmission reduction strategies (e.g., social-distancing, community lockdowns, use of face masks in public, contact tracing etc.) implemented in almost every country or community that is hard-hit with the COVID-19 pandemic, opting, instead, to allow individuals to acquire infection and, hopefully, recover from it. Although some level of this *natural herd immunity* has been achieved in Sweden, the COVID-induced mortality recorded in Sweden far exceed those recorded for neighbouring Scandinavian countries that implemented the aforementioned non-pharmaceutical interventions [32].

In this section, a theoretical condition for achieving community-wide vaccine-induced herd immunity is derived. Let *f_v_* be the fraction of unvaccinated susceptible individuals that have been vaccinated at the disease-free steady-state. That is,

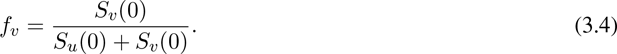

Further, let

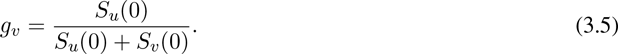

It follows from (3.1), (3.4) and (3.5) that the control reproduction number (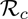) can then be re-written, in terms of *f_v_* and *g_v_*, as follows:

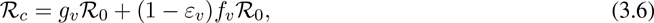

which can be simplified as

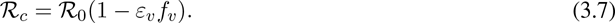

Setting 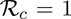 in (3.7), and solving for fv, gives the following vaccine-induced herd immunity threshold for the model (2.1):

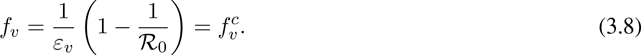

It follows from (3.7) and (3.8) that 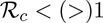 whenever 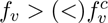. Hence, we achieved the following results:

#### Theorem 3.2.

Consider the model (2.1). The use of an imperfect anti-COVID-19 vaccine will have the following

i. positive population-level impact 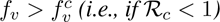.
ii. negative population-level impact if 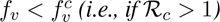.
iii. no population-level impact if 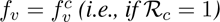.

The result of Theorem 3.2 is numerically-illustrated in Figure 4, showing an increase in the cumulative COVID-induced mortality when 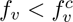 and a decrease when 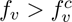. In order to compute the community herd immunity thresholds for the US, New York state and Florida state, we compute the respective values of 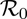, using (3.8) with the baseline values of the community contact rates parameters, *β_a_* and *β_s_*, reduced by 30% (to account for the effect of the implementation of effective social-distancing measures [4]). That is, we set *β_a_* = 0:8400; *β_s_* = 0:4971 for US, *β_a_* = 0:5648; *β_s_* = 0:3871 for New York state and *β_a_* = 0:7675; *β_s_* = 0:4621 for the state of Florida in Equation (3.8) to obtain the values of the basic reproduction number (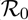) for the US, New York state and the state of Florida. This gives 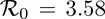 3.04 and 3.12, respectively. Substituting these values of 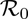 (for each of the three populations) in Equation (3.7), and noting that the vaccine efficacy is assumed to be 80% (i.e., *ε_v_* = 0:8), shows that the respective vaccine-derived community herd immunity thresholds for the entire US, the state of New York and the state of Florida are 90%, 84% and 85%, respectively (as tabulated in Table 4). The epidemiological implication of these analyses is that the pandemic can be effectively controlled and/or eliminated in the US and the states of New York and Florida if at least 90%, 84% and 85% of the respective unvaccinated susceptible populations are vaccinated (using the hypothetical imperfect vaccine with the assumed protective efficacy of 80%). It should be mentioned that these vaccine-derived community herd immunity percentages significantly decrease if the vaccination program is complemented with a public face mask use strategy. In particular, Table 4 shows that the herd immunity threshold for the US reduced to about 78% if half the US population wears mask in public. Face mask usage with compliance of 30% to 50% decreases the herd immunity thresholds for the states of New York and Florida to between 70% to 78%. These are reasonably attainable in the two states. Thus, these simulations show that combining a vaccination program (using the hypothetical anti-COVID-19 vaccine with assumed efficacy of 80%) with a public mask use strategy significantly enhances the prospect of COVID-19 elimination in the states of New York and Florida, and the in entire US nation (albeit elimination is easier to achieve in either of the two states than in the entire US nation).

**Figure 4:**
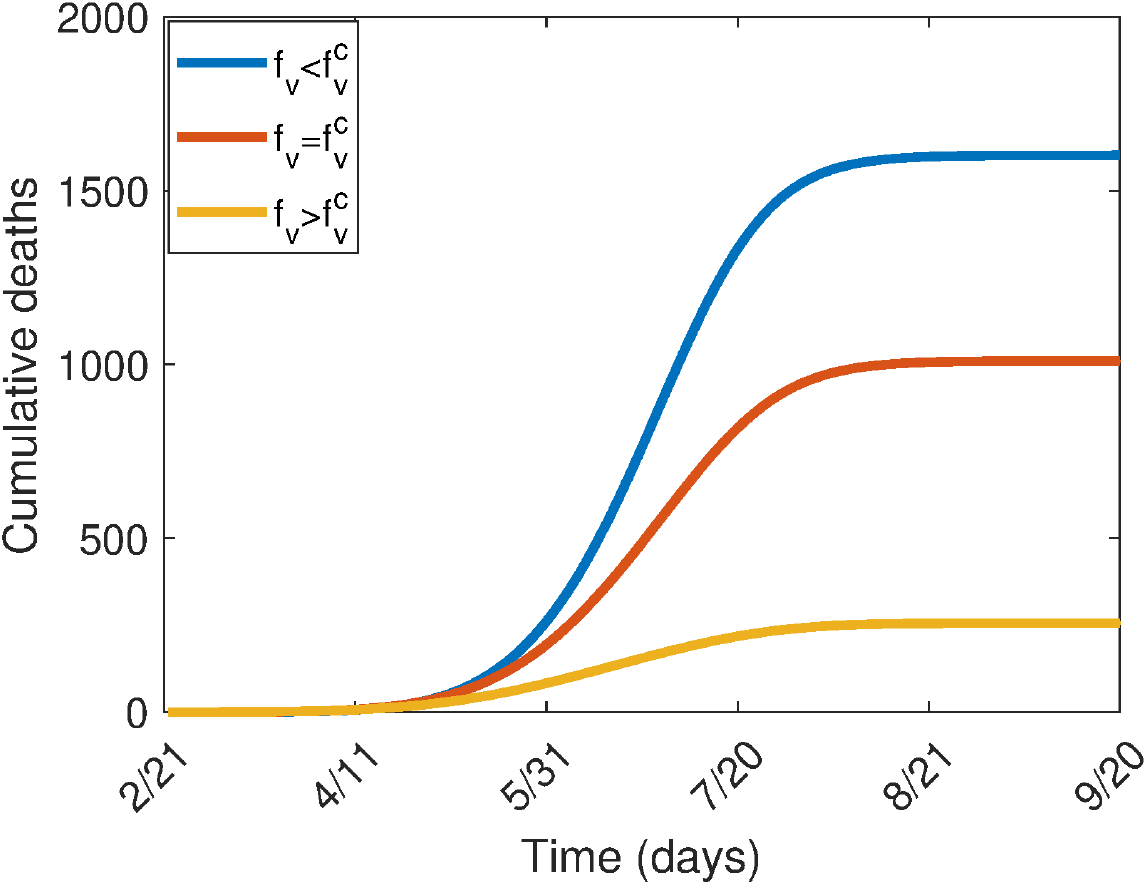
Simulations of the model (2.1), showing the cumulative number of deaths for the state of New York as a function of time. (a) 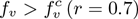 (b) 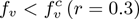 (c) 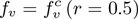. Other parameter values used are as given in Table 3.

**Table 4:**
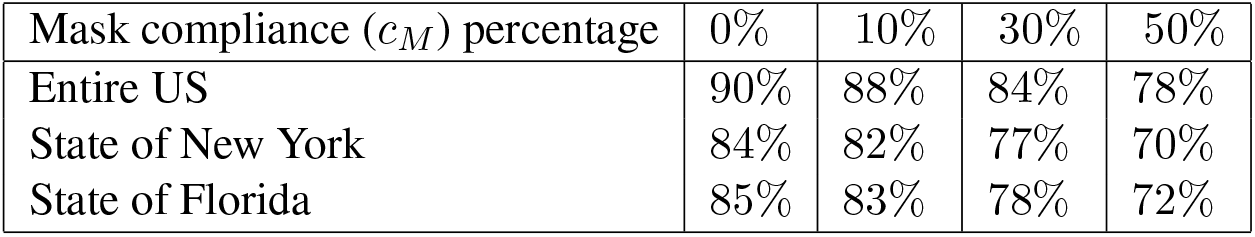
Community herd immunity thresholds for the US, state of New York and state of Florida for the model (2.1). Parameter values are as given in Table 3 with *β_a_* = 0:8400; *β_s_* = 0:4971 for US, *β_a_* = 0:5648; *β_s_* = 0:3871 for New York and *β_a_* = 0:7675; *β_s_* = 0:4621 for Florida (corresponding to an assumed 30% reduction in the baseline value of *β_a_* and *β_s_*, to account for the effect of the implementation of social-distancing measures).

## 4 Numerical Simulations of Vaccine Impact

The model (2.1) is now simulated to assess the population-level impact of the hypothetical imperfect anti-COVID-19 vaccine in the entire US and the US states of New York and Florida. The population-level impact of the vaccination rate (*ξ_v_*) on the burden of the pandemic is monitored first of all. In these simulations, the public face mask use strategy is not implemented. The model (2.1) is then simulated using the baseline parameter values in Table 3 (with *c_M_* = *ε_M_* = 0) and various values of *ξ_v_*. The results obtained, depicted in Figure 5, show that, for the worst-case scenario without vaccination and public mask usage (i.e., the model (2.1) with *ξ_v_* = *S_v_* = *ε_M_* = *c_M_* = 0), the US could record up to 4,000 daily deaths at the pandemic peak (Figure 5 (a)), while the states of New York (Figure 5 (c)) and Florida (Figure 5 (e)) could record up to 2,000 and 1,500 daily deaths at the pandemic peaks, respectively. The corresponding projected cumulative mortality for the entire US, the state of New York and the state of Florida, for the period February 21, 2020 to September 20, 2020, are, 120,000, 66,473 and 47,000, respectively. These simulations show a dramatic reduction in the daily and the cumulative COVID-induced mortality (in comparison to the worst-case scenario without vaccination and mask usage) with increasing values of the vaccination rate *ξ_v_* (from its baseline value). In particular, a 10-fold increase in the baseline value of *ξ_v_* for the

**Figure 5:**
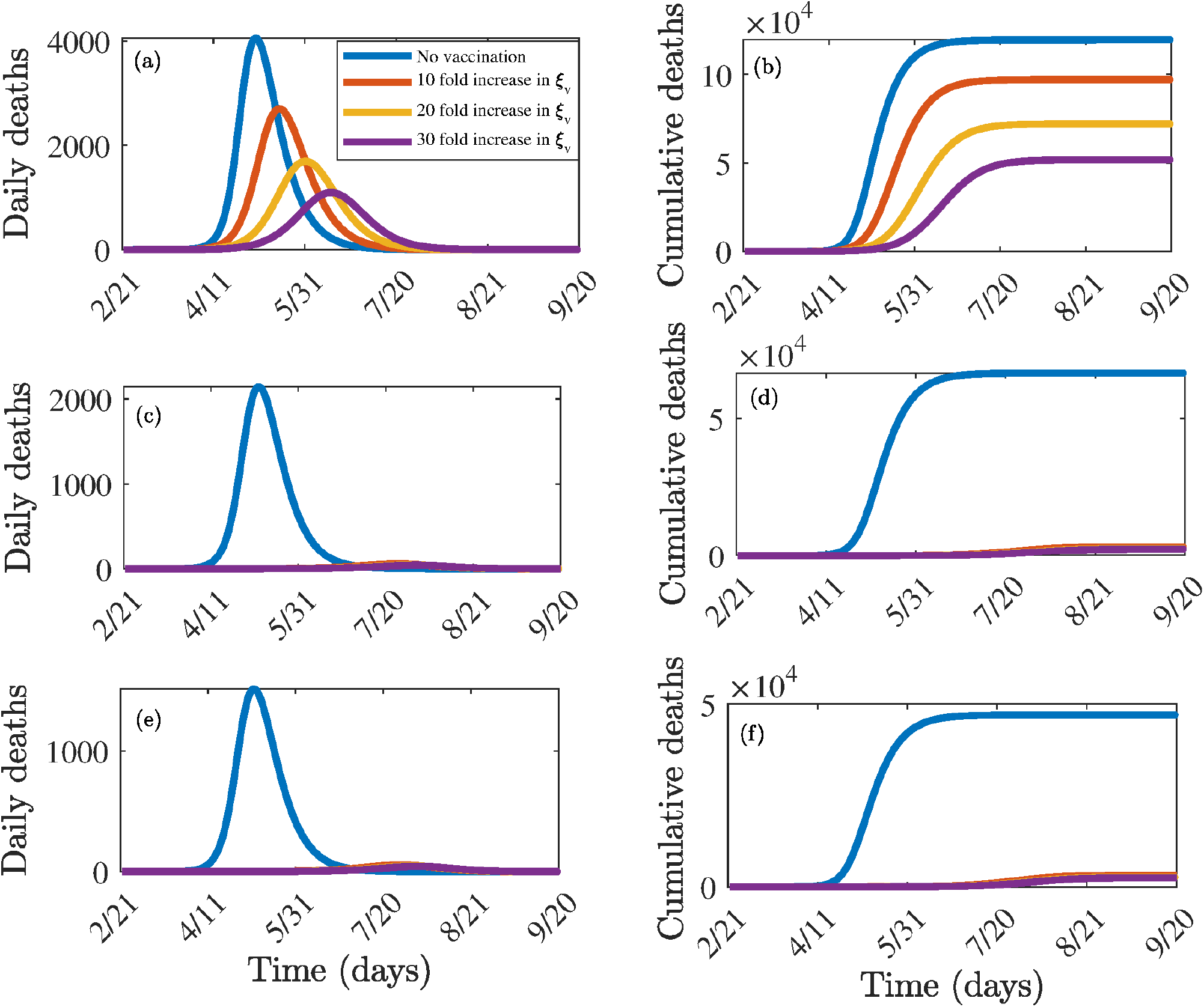
Effect of vaccination coverage. Simulations of the model (2.1) showing the effect of vaccination coverage (*ξ_v_* on COVID-19 burden. (a) Daily deaths for the US. (b) Cumulative deaths for the US. (c) Daily deaths for New York state. (d) Cumulative deaths for New York state. (e) Daily deaths for the state of Florida. (f) Cumulative deaths for the state of Florida. Parameter values used are as given in Table 3, with *cm =ε_m_* =0 and various values of *ξv*.

US resulted in a decrease in the daily number of deaths at the pandemic peak from 4,000 to 2,800, representing a 30% reduction (Figure 5 (a)). Furthermore, this corresponds to a decrease in the projected cumulative mortality from 120,000 to 98,000.

Similarly, a 30-fold increase in the baseline value of *ξ_v_* resulted in a marked reduction of the daily deaths at the pandemic peak to 800 (corresponding to a decrease in the projected cumulative mortality from 120,000 to 50,000). It is evident from Figure 5 that a 10-fold increase in the baseline value of the vaccination rate (*ξ_v_*) for the states of New York (Figures 5 (c) and (d)) and Florida (Figures 5 (e) and (f)) will bring the respective daily deaths at the peak and the projected cumulative mortality to very low levels (i.e., low levels that essentially imply the effective control or elimination of the pandemic). Thus, these simulations suggest that, while a significantly large increase in vaccination rate (from baseline) is necessarily needed to eliminate COVID-19 from the entire US, only a small increase (as low as 10%) in the baseline vaccination rate will be needed to eliminate COVID-19 in the states of New York and Florida. These simulations further show that the prospect of disease elimination using the vaccine (with the assumed protective efficacy of 80%) is more promising in the states of New York and Florida than nationwide (where much larger vaccination coverage will be needed to bring, and maintain, the control reproduction number, 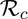, to a value less than unity).

Simulations were also carried out to assess the impact of vaccine efficacy and coverage on this disease dynamics. This is done for the special case of the model in the absence of the use of masks in the public (i.e., the model (2.1) with *c_M_ = ε_m_* = 0). Specifically, a contour plot of the control reproduction number (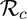) of the model, as a function of coverage (*f_v_*) and vaccine efficacy (*ε_v_*), is depicted in Figure 6. This figure shows a decrease in the value of the reproduction number with increasing coverage and efficacy of the vaccine, as expected. For the assumed 80% efficacy of the hypothetical vaccine (i.e., *ε_v_ =* 0.8) in Table 3 (a), the simulations depicted in Figure 6 (a) show that nationwide elimination of COVID-19 can be achieved (i.e., 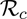 can be brought to a value below unity) if at least 90% of the US populace is vaccinated. For the state of New York, it is shown that such elimination can also occur if the state-wide vaccination coverage is at least 84% (Figure 6 (b)). Similarly, Figure 6 (c) shows that the hypothetical anti-COVID-19 vaccine can lead to the elimination of the pandemic from the state of Florida if at least 85% of the residents of the state are vaccinated.

**Figure 6:**
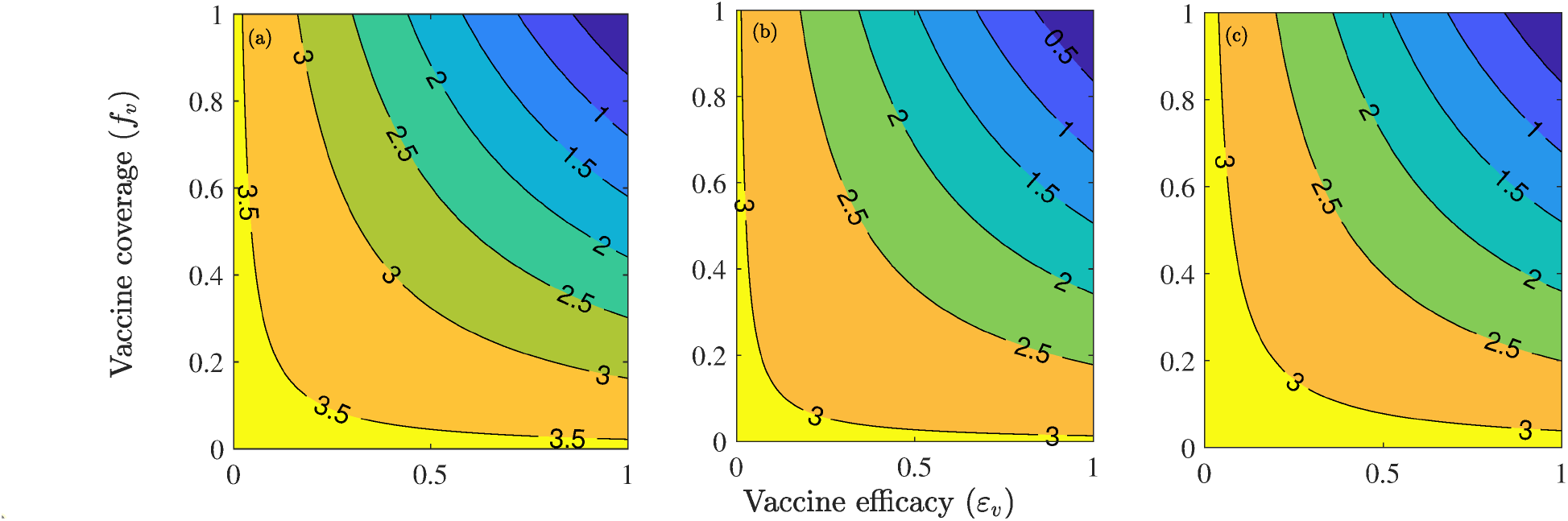
Effects of vaccination as the only intervention strategy. Contour plots of the control reproduction number (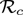) of the model (2.1), as a function of vaccine coverage (*f_v_*) and vaccine efficacy (ε*_v_*), for the baseline scenarios in Table ?? and in the absence of mask usage in public. (a) the entire US (b) the US state of New York (c) the US state of Florida. Parameter values used are as given in Table 3 with *ε_m_ = c_m_ =* 0.

The effect of the combined implementation of vaccination and social-distancing strategies, on the control reproduction number of the model (2.1), is also monitored (for the US and the two US states of New York and Florida) by generating contour plots of 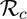, as a function of vaccine coverage and efficacy, for three levels (low, moderate and high) of social-distancing effectiveness. The results obtained, depicted in Figure 7, show that COVID-19 elimination (measured in terms of bringing 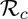 to a value less than unity) is feasible even if social-distancing is implemented at a low level of effectiveness (left panels of Figure 7). In particular, Figure 7 (a) shows that, with the assumed 80% efficacy of the vaccine, the implementation of social-distancing at low effectiveness level will lead to the elimination of COVID-19 in the US if the nationwide vaccination coverage is at least 88%. This vaccination coverage needed to achieve such elimination decreases with increasing effectiveness of the social-distancing measures (Figures 7 (b) and (c)). For the state of New York, for instance, vaccination program combined with social-distancing at low effectiveness level will lead to COVID-19 elimination if at least 80% of the residents of the state are vaccinated (Figure 7 (b)). The vaccination coverage needed for elimination decrease to 67% and 61%, respectively, if the social-distancing measures are implemented at moderate (Figure 7 (e)) and high (Figure 7 (f)) effectiveness levels. Finally, for the state of Florida, combining vaccination with social-distancing at low effectiveness level will lead to COVID-19 elimination in the state if at least 80% of the residents of the state are vaccinated (Figure 7 (g)). The coverage needed for COVID-19 elimination in Florida decrease to 70% and 47%, respectively, for moderate (Figure 7 (h)) and high (Figure 7 (i)) effectiveness levels of the social-distancing strategy. Additional numerical simulations were carried out to assess the community-wide impact of the combined impact of vaccination with a public mask use strategy (as measured in terms of reduction in the value of control reproduction number, 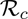). The results obtained, depicted in Figure 8, show a decrease in 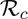 with increasing vaccine efficacy and compliance. For the fixed vaccine efficacy at 80%, the minimum vaccine coverage needed to eliminate COVID-19 in the entire US if public mask use compliance is fixed at 10%, 30% and 50% is 90%, 86% and 82%, respectively (Figures 8 (a), (b) and (c)). Similarly, COVID-19 elimination can occur in the state of New York if the corresponding vaccine coverage are 82%, 80% and 71%, respectively (Figures 8 (d), (e) and (f)). The numbers corresponding to elimination in the state of Florida are 86%, 80% and 73%, respectively (Figures 8 (g), (h) and (i)).

**Figure 7:**
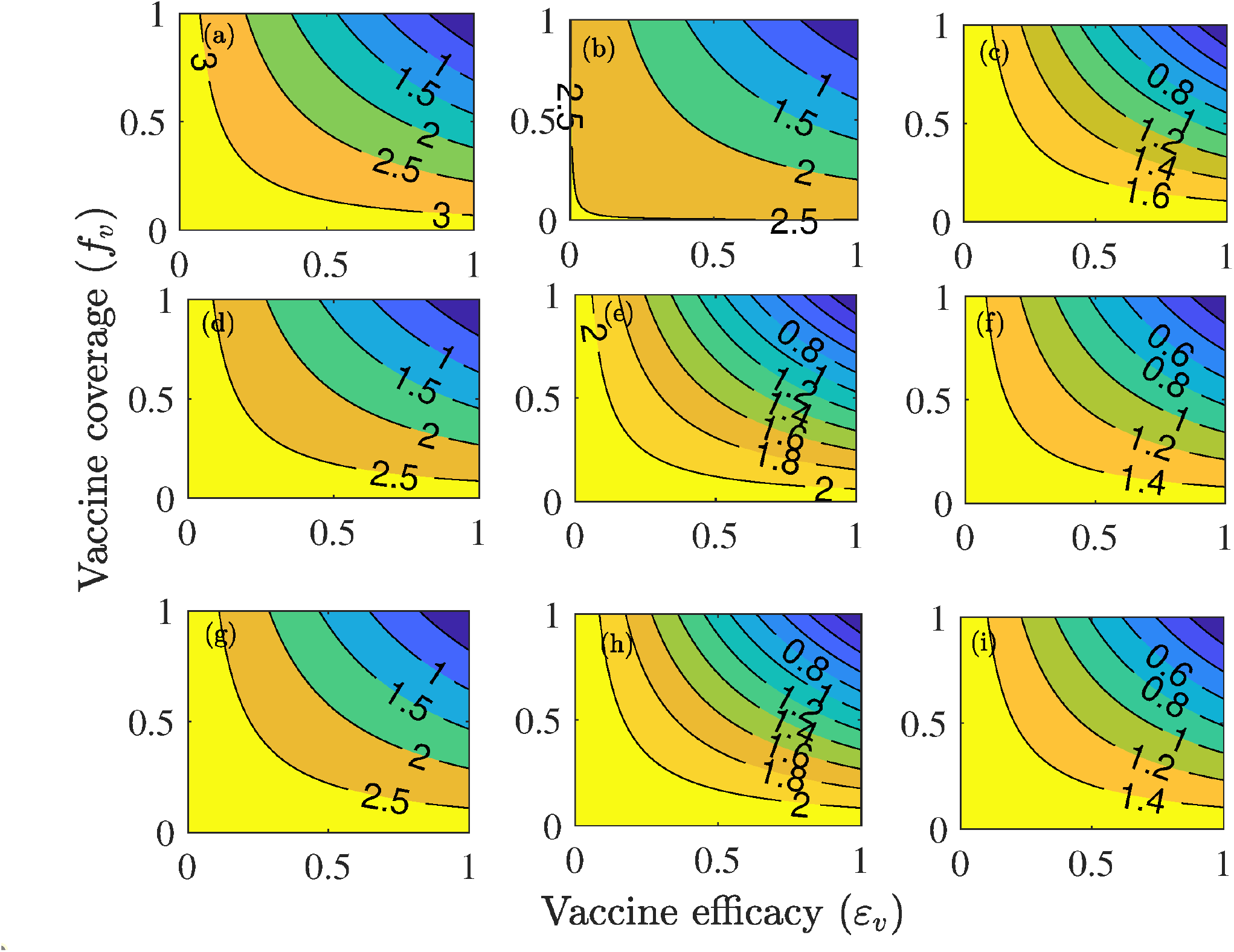
Effect of combined vaccination and social-distancing strategies. Contour plots of the control reproduction number (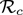), as a function of vaccine coverage (*f_v_*) and vaccine efficacy (*ε_v_*) for the special case of the model (2.1) with no mask usage in the public. (a) Mild-social distancing (10% decrease in *β_s_* and *β_a_*) for the entire US. (b) Moderate social distancing (30% decrease in *β_s_* and *β_a_*) for the entire US. (c) Strict social distancing (50% decrease in *β_s_* and *β_a_*) for the entire US. (d) Mild-social distancing (10% decrease in *β_s_* and *β_a_*) for New York. (e) Moderate social distancing (30% decrease in *β_s_* and *β_a_*) for New York. (f) Strict social distancing (50% decrease in *β_s_* and *β_a_*) for New York. (g) Mild-social distancing (10% decrease in *β_s_* and *β_a_*) for Florida. (h) Moderate social distancing (30% decrease in *β_s_* and *β_a_*) for Florida. (i) Strict social distancing (50% decrease in *β_s_* and *β_a_*) for Florida. Parameter values used are as given in Table 3.

**Figure 8:**
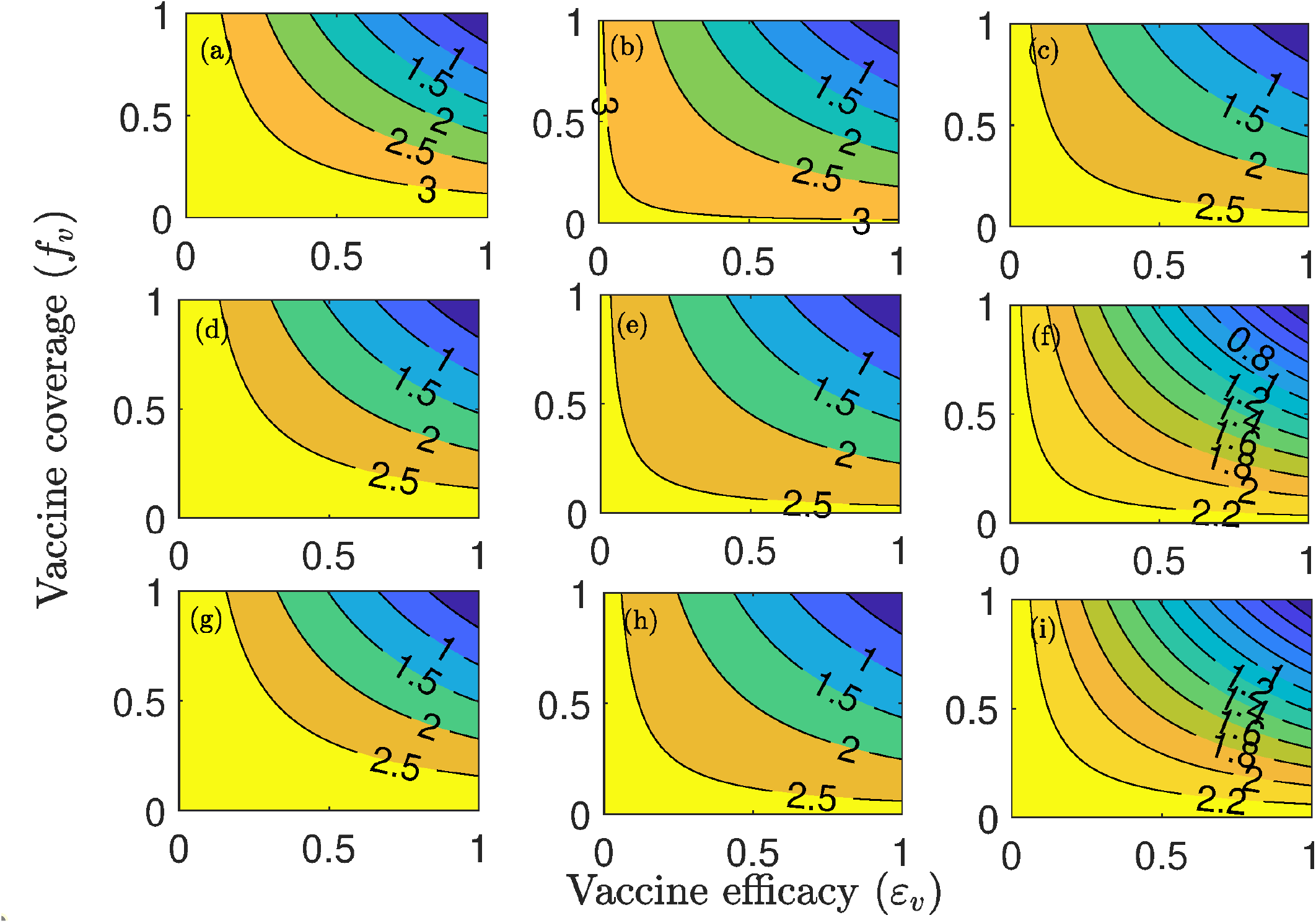
Contour plots of the control reproduction number (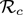), as a function of vaccine coverage (*f_v_*) and vaccine efficacy (*ε_v_*) for various levels of mask compliance. (a) 10% mask compliance for the entire US (b) 30% mask compliance for the entire US (c) 50% mask compliance for the entire US (d) 10% mask compliance for the state of New York (e)30% mask compliance for the state of New York (f) 50% mask compliance for the state of New York (g) 10% mask compliance for the state of Florida (h) 30% mask compliance for the state of Florida (i) 50% mask compliance for the state of Florida. Parameter values used are as given in Table 3 with various values of *c_M_*.

Thus, the simulations in Figures 7 and 8 also emphasize the fact that the prospect of COVID-19 elimination using a vaccine is greatly enhanced if the vaccination program is complemented with another public health intervention (such as social-distancing or the use of face masks in public), particularly if the complementary intervention is implemented at moderate or high effectiveness level.

## 5 Discussion and Conclusions

A novel Coronavirus (COVID-19) emerged in China in December 2019. The virus, which is caused by SARS-CoV-2, rapidly spread to over 210 countries causing over 4 million confirmed cases and 275,000 deaths globally (as of May 8, 2020). Although there are currently no safe and effective vaccine for use in humans, numerous concerted global efforts are underway aimed at developing such vaccine. In fact, a number of candidate vaccines are undergoing advanced stages of clinical trials. One of the most promising of these efforts is the candidate vaccine being developed by a research group at Oxford University, which is expected to be available as early as January 2021 (or, latest, by the spring of 2021). We developed a mathematical model for assessing the the potential community-wide impact of a hypothetical imperfect vaccine against the COVID-19 pandemic in the US. The model we developed, which takes the form of a deterministic system of nonlinear differential equations, was parametrized using COVID-19 data for the entire US, as well as for the US states of New York and Florida. The hypothetical vaccine was assumed to offer imperfect protective efficacy against the acquisition of COVID-19 infection.

The model we developed was rigorously analysed to gain insight into its dynamical features. These analyses reveal that the continuum of disease-free equilibria of the epidemic model is locally-asymptotically stable whenever a certain epidemiological threshold, known as the control reproduction number (denoted by 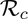), is less than unity. The epidemiological implication of this result is that the COVID-19 pandemic can be effectively controlled, whenever 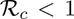, if the initial sizes of the sub-populations of the model are small enough. In other words, for this setting, the routine vaccination against COVID-19 in the US can lead to the effective control of the pandemic if the efficacy and nationwide coverage of the hypothetical vaccine are high enough. In particular, if the vaccine efficacy and coverage are able to bring (and maintain) the reproduction number, 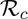, to a value less than unity. Furthermore, threshold quantities for the community vaccine-induced herd immunity were calculated for the entire

US and for the states of New York and Florida. We computed these thresholds for the case where social (physical-distancing is also implemented, and that the social-distancing is implemented at an effectiveness level that reduces the baseline values of the community contact rate parameters (*β_a_* and *β_s_*) by 30%. Our computations show, using the baseline values of the parameters of the model, that at least 90% of the entire US population needs to be vaccinated in order to achieve nationwide vaccine-derived herd immunity. Similarly, at least 84% and 85% of the total susceptible population in the states of New York and Florida, respectively, need to be vaccinated to achieve such herd immunity in the states. These vaccine coverage numbers are certainly on the high side, and may not be easily attained for so many reasons (ranging from the expected insufficient stockpile of the vaccine during the early stages of their deployment, general apathy towards vaccination and the sizable percentage of people that cannot be vaccinated for medical, personal and other reasons). Our simulations showed that if the vaccination strategy is complemented with other public health interventions, such as a public mask use strategy for instance, the minimum herd immunity threshold required to effectively control the COVID-19 pandemic significantly reduces to a more realistically-attainable level. For example, if vaccination (using a vaccine with the assumed efficacy of 80%) is combined with a public face mask strategy with 30% mask compliance nationwide, the herd immunity threshold for the entire US decreases from the 90% (for the mask-free scenario described above) to 78%. Thus, the prospect of the effective control of COVID-19 using routine vaccination is enhanced if it is combined with a public mass use strategy (especially if the effectiveness and coverage of the mask use strategy are high enough).

We also carried out numerical simulations to measure the population-level impact of the rate at which the hypothetical vaccination is administered. The results obtained showed that the COVID-19 burden (as measured in terms of daily and cumulative COVID-induced mortality) decreases with increasing vaccination rate, as expected. Our simulations showed that, while significantly high increases in the vaccination rate (from its baseline value) was necessarily needed to achieve significant reduction in disease burden in the entire US, a relatively small increase in the vaccination rate from its baseline value (as low as 10% increase) will be sufficient to lead to the effective control (or even elimination) of COVID-19 pandemic in the states of New York and Florida. In other words, the prospects of eliminating COVID-19, using the hypothetical vaccine, are more promising in the states of New York and Florida, than nationwide (owing to the unrealistically high vaccine coverage needed to curtail or eliminate the pandemic in the latter).

We also simulated the combined effect of implementing the routine vaccination program with social-distancing (in the absence of face mask usage). We showed that COVID-19 can be effectively controlled using the two interventions even if the effectiveness level of social-distancing is low (as long as the vaccine coverage is high). In particular, our simulations showed that the control reproduction number (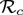) can be reduced to a value less than unity (needed for the effective control of the disease, in line with the local-asymptotic stability property of the continuum of disease-free equilibria, established in Section 3). For example, we showed that for low, moderate and high effectiveness levels for the social-distancing strategy implemented in the state of New York, COVID-19 elimination is feasible if the vaccine coverage is at least 80%, 67% and 61%, respectively (the corresponding figures for the US state of Florida are 80%, 70% and 47%). Thus, these figures suggest that COVID-19 elimination (using the combined vaccination and social-distancing strategy) is more likely in the US state of Florida, followed by the US of state of New York and then nationwide.

We assessed the population-level impact of combining the routine vaccination strategy with a public mask use strategy. Our simulations show a decrease in the burden of the COVID-19 pandemic (again as measured in terms of reduction in the value of the reproduction number 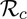) with increasing efficacies and compliance of the vaccine and mask usage. It was shown that, for the case of the entire US, if the vaccine efficacy is fixed at the assumed 80% and mask compliance is fixed at the low value of 10%, COVID-19 can be eliminated in the US if the vaccine coverage is at least 90%. The vaccine coverage decreases to 86% and 82%, respectively, if mask use compliance is increased to 30% and 50% nationwide (the numbers for US state of New York, corresponding to the aforementioned mask use coverage are 82%, 80% and 71%, respectively; similarly, the numbers for the US state of Florida are 86%, 80% and 73%, respectively). Again, these result confirmed that COVID-19 elimination is more feasible is vaccination is combined with another intervention (mask use in public, in this case), and that such elimination is more easily attainable in the US states of New York and Florida than nationwide.

In summary, this study shows that the prospect of effectively controlling COVID-19 in the states of New York and Florida, as well as nationwide, is promising using the hypothetical imperfect anti-COVID-19 vaccine. The prospect of such effective control and/or elimination is greatly enhanced if vaccination is combined with other interventions that limit community transmission, such as social-distancing and mask use in public. Furthermore, elimination is more likely to be achieved using a vaccine in the states of New York and Florida than nationwide (since the latter requires much higher, and probably realistically-unattainable, vaccination coverage to achieve elimination).

## Data Availability

All data source were indicated

## Acknowledgments

One of the authors (ABG) acknowledge the support, in part, of the Simons Foundation (Award #585022) and the National Science Foundation (Award #1917512). CNN acknowledges the support of the Simons Foundation (Award #627346).

## References

[1] World Health Organization, “Coronavirus disease (COVID-19) technical guidance,” WHO (Accessed on March 4, 2020). Online Version https://www.who.int/emergencies/diseases/novel-coronavirus-2019/technical-guidance

[2] World Health Organization, “Emergencies, preparedness, response. Pneumonia of unknown origin -China,” Disease Outbreak News (5 January, Accessed on March 5, 2020). Online Version https://www.who.int/csr/don/05-january-2020-pneumonia-of-unkown-cause-china/en/

[3] Q. Li, X. Guan, P. Wu, X. Wang, L. Zhou, Y. Tong, R. Ren, K. S. Leung, E. H. Lau, J. Y. Wong, et al., “Early transmission dynamics in Wuhan, China, of novel coronavirus-infected pneumonia,” New England Journal of Medicine (2020).

[4] C. N. Ngonghala, E. Iboi, S. Eikenberry, M. Scotch, C. R. MacIntyre, M. H. Bonds, and A. B. Gumel, “Mathematical assessment of the impact of non-pharmaceutical interventions on curtailing the 2019 novel coronavirus,” Mathematical Biosciences. 325, 108364 (2020).

[5] “Center for Systems Science and Engineering at Johns Hopkins University. COVID-19,” (2020). Online Version https://github.com/CSSEGISandData/COVID-19

[6] S. Knvul and T. Katie, “More coronavirus vaccines and treatments move toward human trials,” New York Times (Assessed on May 2, 2020). Online Version https://www.nytimes.com/2020/04/08/health/coronavirus-vaccines.html

[7] World Health Organization, “Coronavirus disease 2019 (COVID-19): situation report, 46,” WHO (2020).

[8] S. Matthews, “U.S. Jobless Rate May Soar to 30%.” Bloomberg Report (May 2, 2020). Online Version https://www.bloomberg.com/news/articles/2020-03-22/fed-s-bullard-says-u-s-jobless-rate-may-soar-to-30-in-2q

[9] D. Rotman, “Stop COVID or save the economy? We can do both.” MIT Technology Review. (May 2, 2020). Online Version https://www.technologyreview.com/2020/04/08/998785/stop-covid-or-save-the-economy-we-can-do-both

[10] N. M. Ferguson, D. Laydon, G. Nedjati-Gilani, N. Imai, K. Ainslie, M. Baguelin, S. Bhatia, A. Boonyasiri, Z. Cucunubá, G. Cuomo-Dannenburg, et al., “Impact of non-pharmaceutical interventions (NPIs) to reduce COVID-19 mortality and healthcare demand,” London: Imperial College COVID-19 Response Team, March 16 (2020).

[11] S. E. Eikenberry, M. Muncuso, E. Iboi, T. Phan, E. Kostelich, Y. Kuang, and A. B. Gumel, “To mask or not to mask: Modeling the potential for face mask use by the general public to curtail the covid-19 pandemic,” Infectious Disease Modeling 5, 293-308 (2020).

[12] E. Callaway, “Scores of coronavirus vaccines are in competition — how will scientists choose the best?” a natureresearch journal (Assessed on April 30, 2020).

[13] D. Kirkpatrick, “In race for a Coronavirus vaccine, an Oxford Group Leaps Ahead,” New York Times (May 2, 2020). Online Version https://www.nytimes.com/2020/04/27/world/europe/coronavirus-vaccine-update-oxford.html

[14] K. Mizumoto and G. Chowell, “Transmission potential of the novel coronavirus (COVID-19) onboard the Diamond Princess Cruises Ship, 2020,” Infectious Disease Modelling (2020).

[15] J. Hellewell, S. Abbott, A. Gimma, N. I. Bosse, C. I. Jarvis, T. W. Russell, J. D. Munday, A. J. Kucharski, W. J. Edmunds, F. Sun, et al., “Feasibility of controlling COVID-19 outbreaks by isolation of cases and contacts,” The Lancet Global Health (2020).

[16] A. J. Kucharski, T. W. Russell, C. Diamond, Y. Liu, J. Edmunds, S. Funk, R. M. Eggo, F. Sun, M. Jit, J. D. Munday, et al., “Early dynamics of transmission and control of COVID-19: a mathematical modelling study,” The Lancet Infectious Diseases (2020).

[17] P. van den Driessche and J. Watmough, “Reproduction numbers and sub-threshold endemic equilibria for compartmental models of disease transmission,” Mathematical Biosciences 180, 29-48 (2002).

[18] O. Diekmann, J. A. P. Heesterbeek, and J. A. Metz, “On the definition and the computation of the basic reproduction ratio *R*_0_ in models for infectious diseases in heterogeneous populations,” Journal of Mathematical Biology 28, 365-382 (1990).

[19] E. Dong, H. Du, and L. Gardner, “An interactive web-based dashboard to track COVID-19 in real time,” The Lancet Infectious Diseases (2020).

[20] E. Dong, H. Du, and L. Gardner, “Coronavirus COVID-19 Global Cases by Johns Hopkins CSSE,” The Lancet Infectious Diseases (2020). Online Version https://gisanddata.maps.arcgis.com/apps/opsdashboard/index.html#/bda7594740fd40299423467b48e9ecf6

[21] World Health Organization, “Coronavirus disease (COVID-2019) situation reports,” WHO (Accessed on March 19, 2020). Online Version https://www.who.int/emergencies/diseases/novel-coronavirus-2019/situation-reports

[22] Centers for Disease Control and Prevention, “Coronavirus disease 2019 (COVID-19),” National Center for Immunization and Respiratory Diseases (NCIRD), Division of Viral Diseases (Accessed on March 4, 2020). Online Version https://www.cdc.gov/coronavirus/2019-ncov/index.html

[23] A. Davies, K.-A. Thompson, K. Giri, G. Kafatos, J. Walker, and A. Bennett, “Testing the efficacy of homemade masks: would they protect in an influenza pandemic?” Disaster Medicine and Public Health Preparedness 7, 413–418(2013).

[24] S. A. Lauer, K. H. Grantz, Q. Bi, F. K. Jones, Q. Zheng, H. R. Meredith, A. S. Azman, N. G. Reich, and J. Lessler, “The incubation period of coronavirus disease 2019 (covid-19) from publicly reported confirmed cases: estimation and application,” Annals of internal medicine (2020).

[25] R. Li, S. Pei, B. Chen, Y. Song, T. Zhang, W. Yang, and J. Shaman, “Substantial undocumented infection facilitates the rapid dissemination of novel coronavirus (SARS-CoV2),” Science (2020).

[26] R. Verity, L. C. Okell, I. Dorigatti, P. Winskill, C. Whittaker, N. Imai, G. Cuomo-Dannenburg, H. Thompson, P. G. Walker, H. Fu, et al., “Estimates of the severity of coronavirus disease 2019: a model-based analysis,” The Lancet Infectious Diseases (2020).

[27] L. F. Moriarty, “Public health responses to covid-19 outbreaks on cruise ships—worldwide, February-March 2020,” MMWR. Morbidity and Mortality Weekly Report 69 (2020).

[28] F. Zhou, T. Yu, R. Du, G. Fan, Y. Liu, Z. Liu, J. Xiang, Y. Wang, B. Song, X. Gu, et al., “Clinical course and risk factors for mortality of adult inpatients with COVID-19 in Wuhan, China: a retrospective cohort study,” The Lancet (2020).

[29] B. Tang, X. Wang, Q. Li, N. L. Bragazzi, S. Tang, Y. Xiao, and J. Wu, “Estimation of the transmission risk of the 2019-nCoV and its implication for public health interventions,” Journal of Clinical Medicine 9, 462 (2020).

[30] R. M. Anderson and R. M. May, “Vaccination and herd immunity to infectious diseases,” Nature 318, 323-329 (1985).

[31] R. M. Anderson, “The concept of herd immunity and the design of community-based immunization programmes,” Vaccine 10, 928-935 (1992).

[32] T. L. Friedman, “Is Sweden doing it right?” New York Times (Assessed on May 4, 2020). Online Version https://www.nytimes.com/2020/04/28/opinion/coronavirus-sweden.html

